# Understanding the burden and mitigating risks in the utilisation of the Emergency Medical Services in the management of community mental health emergencies

**DOI:** 10.1101/2021.11.28.21266975

**Authors:** Ian Howard, Nicholas Castle, Loua Al Shaikh, Robert Owen

**Author notes:** (Corresponding author) Ian Howard.

## Abstract

**Introduction:** Mental health disorders are highly prevalent globally with access to appropriate care oftentimes problematic. The Emergency Medical Services (EMS) have been suggested as a potential solution to improve access, however, it is unclear whether these services provide the most appropriate response with respect to the needs of patients experiencing a mental health emergency.

**Methods:** A multi-method study was conducted to assess the current burden and potential risks associated with the transportation of patients experiencing a mental health emergency by EMS. Part 1 utilised a cross-sectional study of routinely collected clinical data for the period January 2018 to December 2019. Part 2 employed a systematic risk identification methodology focused on identifying the hazards associated with the transportation of this patient cohort, to identify key action plans towards mitigating their occurrence.

**Results:** Patients experiencing a mental health emergency were transported at an average rate of 11.96 per 1000 transports. Approximately 7% were administered prehospital sedation, with Ketamine administered as the most common sedative. Altogether, 50 potential hazards were identified involving the transportation of patients experiencing a mental health emergency. The *Patient Assessment* subprocess contained the most potential hazards/failure points (n=13). Risks categorized as *occasional* (n=33) and *moderate* (n=16) made up the majority.

**Conclusion:** The burden of mental health emergencies on EMS is considerable, with the potential for several significant risks. Despite this, there was unequal focus on the development of care pathways and clinical guidance for this patient cohort compared with the more “traditional” case types serviced by EMS. Consequently, we identified several strategic-level action plans to mitigate these hazards and improve the delivery of care for these patients in the community.

## INTRODUCTION

Mental health disorders are highly prevalent globally, with approximately 18% of the population meeting criteria for having experienced a common mental health disorder in the previous 12 months, and as many as 29% of the population expected to experience a mental health disorder in their lifetime (1). However, despite this prevalence, access to the appropriate care at the optimal time for individuals who experience mental health-related issues is often complex or otherwise problematic, especially given the significant gap between the demand for and availability of mental health services (2,3). Furthermore, these barriers to access of care are particularly prominent amongst black and ethnic minority communities (4), women (5) and other at-risk individuals such as troubled youth or the LBGTQ+ community (6,7).

Given their role as a significant entry point into the broader health services, utilisation of the emergency medical services has been suggested as a potential solution to partly improve access to specialised mental health services (8-11). There is, however, limited evidence to indicate that this increase in caseload is likely to be considerable, with the potential to significantly impact service delivery, quality of care, and patient safety (8-11). This is further confounded by concerns that mental health emergencies have not historically constituted what could be considered a “traditional” case type serviced by or exposed to EMS, such as myocardial infarction, stroke, or respiratory emergencies (10). Consequently, it is unclear whether EMS provides the most appropriate response with respect to the clinical needs and outcomes of patients experiencing a mental health emergency.

The treatment and transportation of mental health emergencies in the community is fraught with risks, particularly if carried out by ill-prepared personnel and services. In order to begin to address these risks, it is essential to understand the limitations of EMS in the management of mental health emergencies and identify priorities to mitigate these risks and improve service offering. Consequently, the aim of this study was to further understand the burden exerted by this patient group on emergency service provision and prospectively identify potential risks and hazards in the utilisation of EMS for the transportation of patients experiencing a mental health emergency in the community.

## METHODS

In order to comprehensively address the research aim, a multi-method study was conducted, divided into two parts. Part 1 focused on measuring the current burden of mental health emergencies transported by EMS through a cross-sectional study of service activity for this patient cohort. Part 2 focused on identifying the hazards and risks associated with the transportation of patients suffering a mental health emergency in order to identify key action plans towards mitigating their occurrence and effect and to ultimately improve the service offering for these patients in the community.

### Setting

The study was conducted in the Hamad Medical Corporation Ambulance Service (HMCAS), the government-funded national ambulance service of Qatar, serving a population of approximately 2.7 million people, with a combined daily average call rate of approximately 800 - 900 emergency or primary community cases and 300 non-emergency or inter-facility transfers per day. The service follows a USA/UK/Australia EMS model of independently licenced emergency care practitioners for its frontline staff and operates a two-tiered system of Ambulance Paramedic (AP) staffed ambulances and Critical Care Paramedic (CCP) staffed fast-response vehicles.

As is common in the region, the varied mix of nationalities and cultures that make up the population of Qatar is echoed in the broader healthcare workforce. This has added an extra layer of complexity in the day-to-day operations of delivering healthcare in the region, given the diverse backgrounds in education and experience over and above the ethnic, cultural and language differences amongst healthcare workers in Qatar. Within the ambulance service, CCPs are recruited from EMS organisations around the world with specific training and experience in prehospital emergency care. APs, however, are sourced from a variety of allied healthcare backgrounds and then receive additional in-service bridging training once in-country. Consequently, differences in training and experience with treating patients with mental health emergencies amongst EMS staff is significant.

### Part 1: Cross-sectional study

Part 1 utilised routinely collected clinical data extracted from the HMCAS electronic health record database for the period January 2018 to December 2019. Patients were identified by provisional diagnosis and primarily ascertained by the attending EMS clinician when either the patient or patient’s family positively identified a previous history of a mental health disorder, and for whom no other discernible medical or traumatic reason for the call for service could be identified. Patients <= 18 years old, patients who refused transport, and patients transferred between healthcare facilities were excluded from the analysis.

Data regarding caseload activity was captured and compared between mental health emergencies and case types conventionally serviced by EMS, for which well-defined/established practice guidelines and care pathways already existed within HMCAS. For the purposes of this study, these included acute myocardial infarction (AMI), stroke and asthma. Furthermore, demographic data and key interventions in the treatment and transportation of mental health emergencies were captured to further understand the patient population and their service utilisation. Of particular interest was the subgroup of transported patients who required prehospital sedation, where data regarding drug type, number of doses, and mean dose per drug type was collected. Data for part 1 were analysed using univariate descriptive statistics and reported using frequencies and percentages or mean and standard deviation. A basic dashboard of key patient and service metrics was developed using data from the cross-sectional survey and presented. Data were collated and analysed using Microsoft Excel (Microsoft Office Professional 2008).

### Part 2: Healthcare Failure Mode and Effects Analysis

Part 2 utilised the Healthcare Failure Mode and Effects Analysis (HFMEA) framework, a qualitative-based, systematic technique for risk identification and failure analysis developed by the United States Department of Veteran Affairs National Centre for Patient Safety. The HMFEA is a prospective, multistage, multidisciplinary, modified version of the generic Failure Mode Effects Analysis technique, adapted for the healthcare setting. Throughout the early rounds of the HFMEA, additional input into identifying processes and potential failure points was provided by multiple alternative data sources, including clinical audit data and occurrence-variance-accident reporting system data(11,12).

A multidisciplinary team from within HMCAS was assembled, with the primary focus for inclusion to ensure adequate representation from all departments that contribute towards operational readiness and activity, including operational staff (ground and aeromedical), senior operations managers, as well as representatives from executive management, training, clinical audit, and governance. Multiple rounds of both face-to-face meetings and online correspondence occurred throughout the HFMEA process in order to achieve the study aim.

#### Process and Subprocess Description

For Round 1, an initial process and subprocess map were drafted, through consensus agreement, for what was determined to be a generic EMS call for a patient presenting with a potential mental health emergency. A follow-up draft process map was re-circulated amongst the panel members individually for further reflection and refinement. A final draft was then independently validated by an operational CCP not involved in the process map development or audit.

#### Hazard Identification and Analysis

For round 2, potential hazards and/or points of failure were identified for each sub-process, again by team consensus. For the purposes of this study, a hazard or failure point was defined as “ways or manners in which a sub-process may potentially fail to reach and/or provide its anticipated result or outcome” (12). The draft list was similarly supplemented by data from the audit highlighted above. The updated list was then circulated among team members for independent hazard analysis and subsequent consensus agreement. Consensus was determined when ≥ 70% of participants were in agreement about assigned scores or categories throughout data collection.

For round 3, a hazard analysis was conducted on the hazards and failure points identified during round 2. The HFMEA hazard analysis differs from the traditional FMEA approach in that severity and probability of failure points are assigned a category and not a numerical value (Table 1). Hazard scores were then calculated based on the assigned categories using a matrix to determine a final value (Table 2). An HFMEA decision tree was followed using the final hazard matrix scores to determine criticality, controllability, and detectability of each failure point (see Figure 1 for decision tree and definitions).

**Table 1:** Severity and Probability definitions.

| **Table 1: Severity and Probability definitions** |  |
| --- | --- |
| **Severity Rating Scale Definitions (12)** |  |
| **Catastrophic** | Death or major permanent loss of function (sensory, motor, physiologic, or intellectual), suicide, rape, hemolytic transfusion reaction, surgery/procedure on the wrong patient or wrong body part, infant abduction, or infant discharged to the wrong family |
| **Major** | Permanent lessening of bodily functioning (sensory, motor, physiologic, or intellectual), disfigurement, surgical intervention required, increased length of stay for 3 or more patients, increased level of care for 3 or more patients |
| **Moderate** | Increased length of stay or increased level of care for 1 or 2 patients |
| **Minor** | No injury nor increased length of stay nor increased level of care |
| **Probability Rating Scale Definitions (12)** |  |
| **Frequent** | Likely to occur immediately or within a short period (May happen several times in 1 year) |
| **Occasional** | Probably will occur (May happen several times in 1 to 2 years) |
| **Uncommon** | Possible to occur (May happen sometime in 2 to 5 years) |
| **Remote** | Unlikely to occur (May happen sometime in 5 to 30 years) |

**Table 2:**
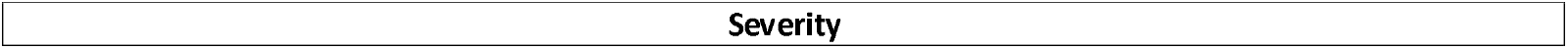
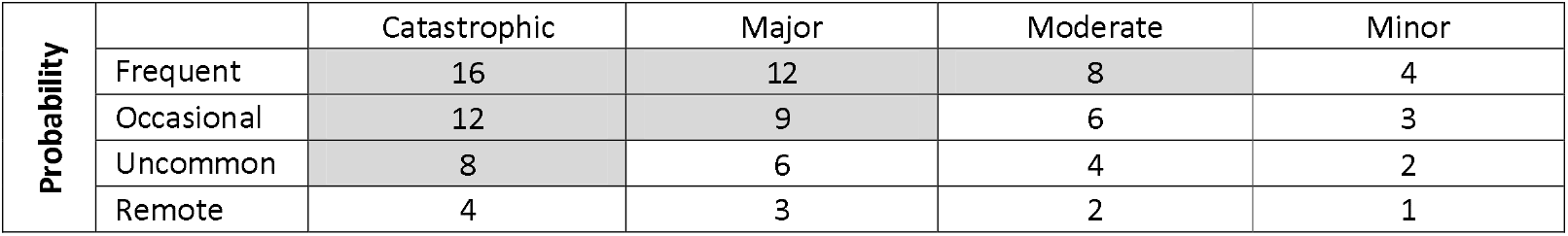
HMFEA Hazard Scoring Matrix (12)

| **Table 2: HFMEA Hazard Scoring Matrix (12)** |
| --- |
| **Severity** |

| Probability |  | Catastrophic | Major | Moderate | Minor |
| --- | --- | --- | --- | --- | --- |
|  | Frequent | 16 | 12 | 8 | 4 |
|  | Occasional | 12 | 9 | 6 | 3 |
|  | Uncommon | 8 | 6 | 4 | 2 |
|  | Remote | 4 | 3 | 2 | 1 |

**Figure 1:**
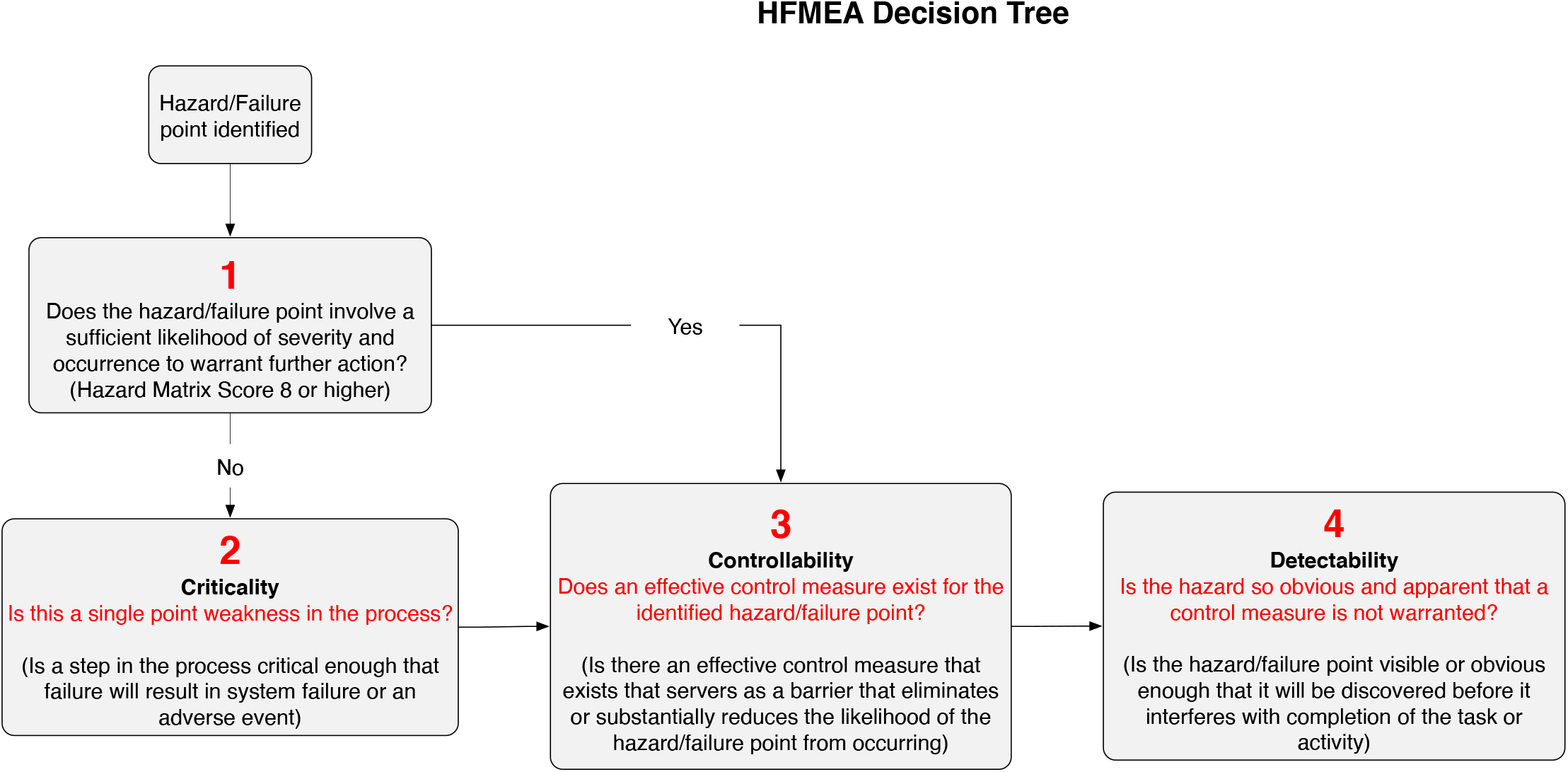
HFMEA decision tree

#### Action Plan Recommendations

A fourth and final consensus round was then conducted to determine whether or not further action was required to mitigate the hazards and failure points identified as priorities, and to identify potential action plans to achieve this. Failure points that scored ≥ 8 following the hazard analysis were automatically included for further discussion for action.

### Institutional Review Board

Ethical approval to conduct the study was granted by the Medical Research Centre of the Hamad Medical Corporation, Qatar (Ref. no.: MRC-01-20-662).

## RESULTS

### Part 1: Cross-sectional study

During the study period, a total of 5484 patients experiencing a mental health emergency were transported, at an average rate of 11.96 per 1000 total patient transports, over twice the rate of either stroke (5.56 per 1000 patient transports) or AMI (5.25 per 1000 patient transports), and second only to asthma cases (14.17 per 1000 patient transports) (Figure 2). Of further interest to note, patients treated and transported for mental health emergencies were generally younger and with a higher proportion of female patients than the standardised population in Qatar (supplementary material 1). Approximately 7% (n = 396) of all patients transported with a mental health emergency had a repeat transport during the study period, with 5% (n = 18) of this group requesting EMS assistance greater than five times over the study period. The highest number of transports of a single patient for a mental health emergency over the 24-month study period was 25.

**Figure 2:**
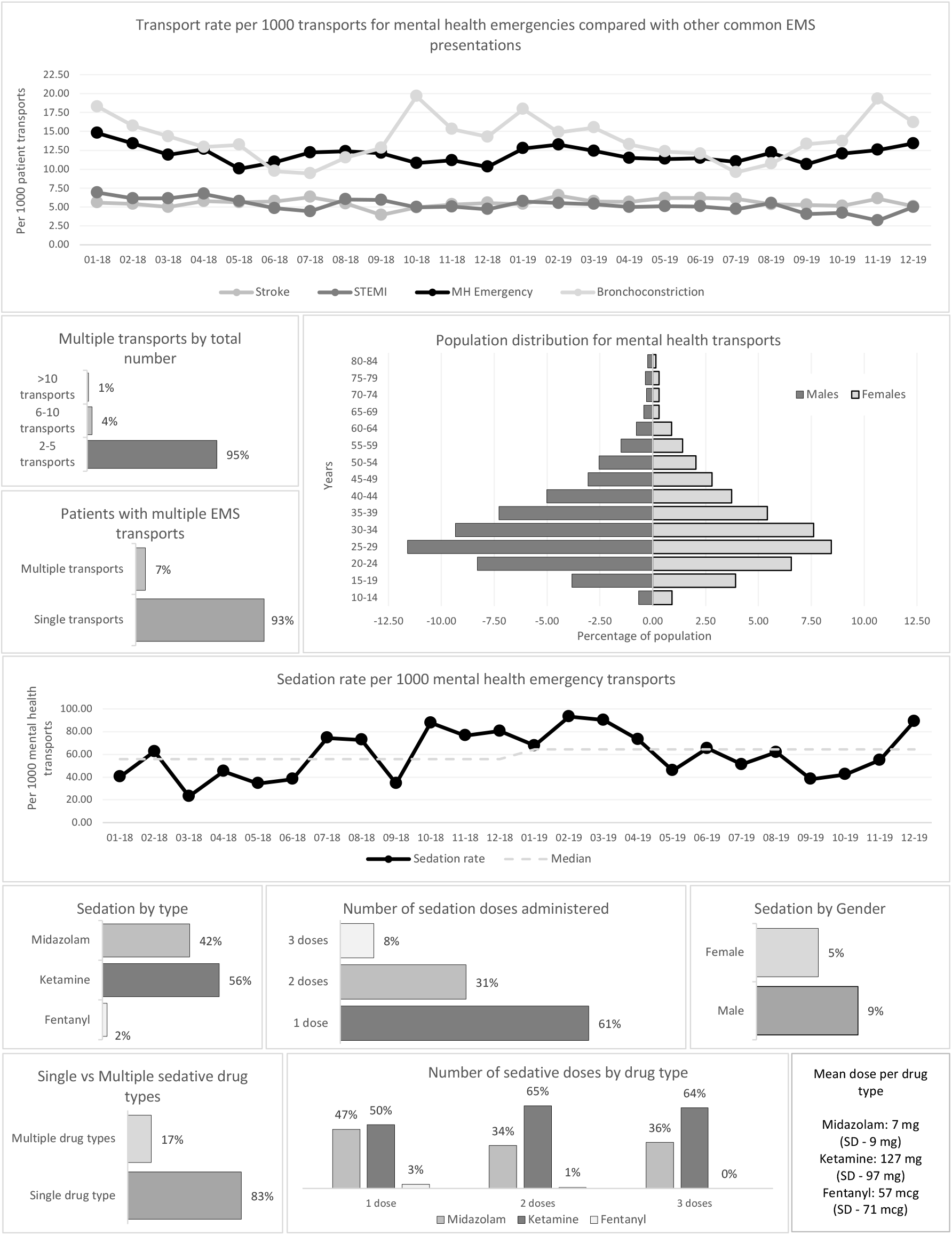
Activity dashboard

Approximately 7% (n = 402) of all mental health emergencies serviced during the study period were administered prehospital sedation prior to arrival at hospital, with a year-on-year increase from 55.82 per 1000 mental health transports in 2018, to 64.39 per 1000 mental health transports in 2019. Male patients were sedated almost twice as often as female patients, with Ketamine being the most commonly administered sedative, given in just over half of cases (56%). Of the cases in which sedation was administered, approximately 17% (n = 54) received two different sedative drug types, and approximately 39% (n = 157) of patients required multiple sedative doses. The mean drug dose administered per patient encounter was 7mg for Midazolam, 127 mg for Ketamine, and 57 mcg for Fentanyl.

### Part 2: Healthcare Failure Mode and Effects Analysis

A process map for what was determined to be a generic EMS call for a patient presenting with a mental health emergency was drafted, consisting of six major processes, including the following (Figure 3):

- Call taking and dispatch
- Patient assessment
- Patient management
- Patient disposition
- Patient safety and quality of care
- General preparedness

**Figure 3:**
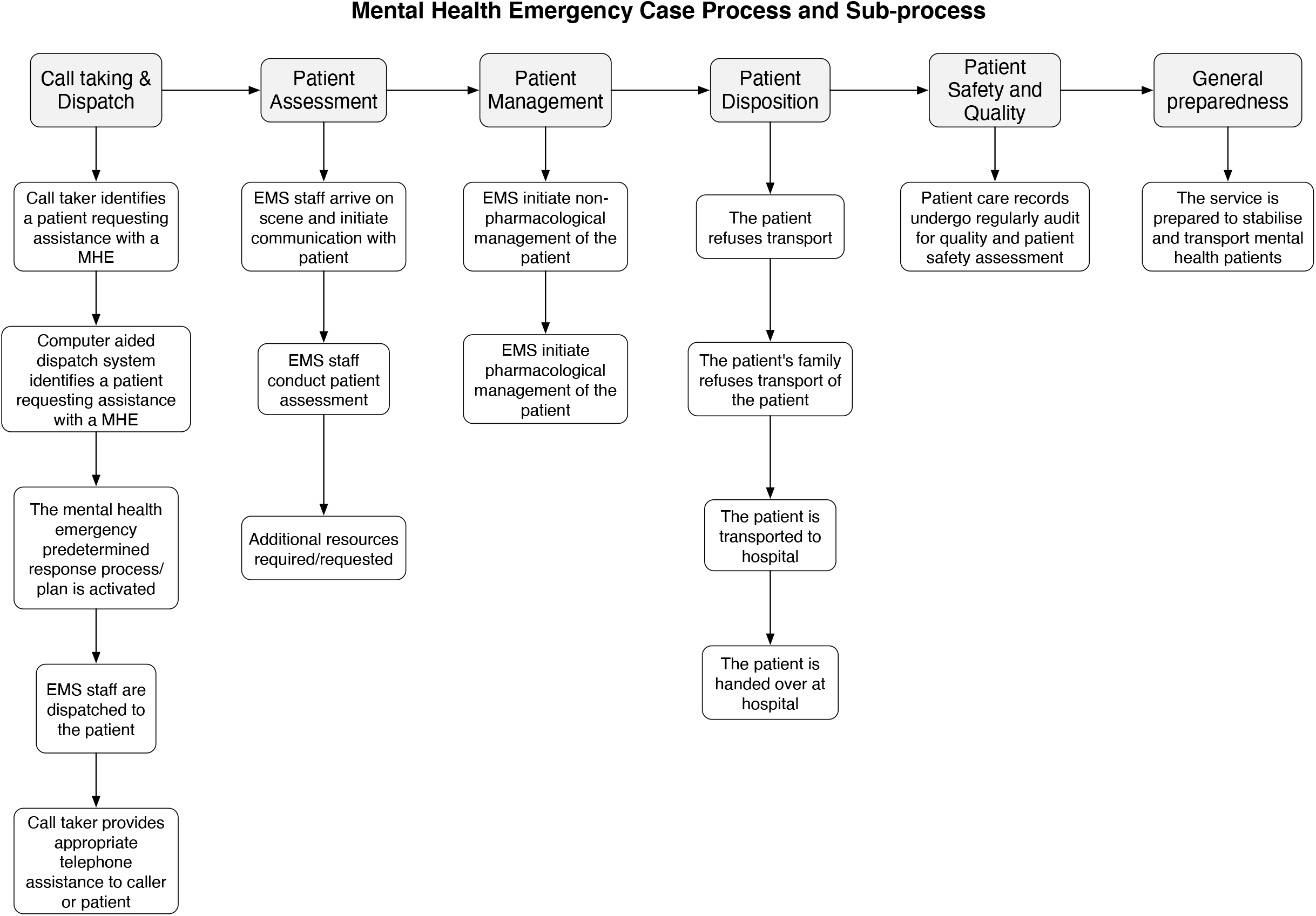
Process/Sub-process map

Among these, 16 separate sub-processes were identified where altogether, 50 potential hazards/ failure points were described amongst the separate sub-processes (Table 3). The *Patient Assessment* subprocess contained the most potential hazards/failure points (n=13, 26%) followed by *Call Taking & Dispatch, Patient Management* and *Patient Disposition*, with 10 hazards each (20%).

### Hazard Analysis

For the probability categorization, the failure points categorized as occasional (n = 33, 66%) made up the majority, followed by uncommon (n = 10, 20%) (Table 3). In terms of severity, failure points categorized as moderate (n = 32, 64%) contributed the largest group, followed by major (n = 16, 32%). Based on the results of the hazard scoring, 11 failure points (22%) scored 8 or higher, thus warranting further discussion for action. An additional 32 failure points (64%) were, through consensus, deemed significant enough to warrant further discussion or which could otherwise be addressed with the outlined action plans for implementation, despite a hazard score lower than 8 (Table 3). No failure points scoring higher than 8 were found to be adequately controlled for and excluded from further action. The remaining 7 (14%) potential failure points described were accepted given either their low likelihood of occurring or deemed to be outside of the influence of service operations.

### Action Plan Recommendations

Given the aim of the study, all actions to be implemented were developed with a strategic focus in mind. The purposes of these actions were from the outset, intended to serve as a starting point in the development of a mental health emergency care pathway within HMCAS and as such were developed with this understanding. Consequently, seven strategic-level action items were identified based on the results of the analysis. These included the following:

- AP (Action plan) 1: Implementation and training of call centre staff on an updated mental health call taking and dispatch protocol. At the time of writing, the computer aided dispatch system used by HMCAS had introduced a focused mental health call taking and dispatch code as part of the latest software update and was used as the basis for this action plan
- AP 2: Development and implementation of dedicated, female staffed, response units. Given the high proportion of female patients that made up the mental health emergency cohort observed in the 24-month study period, the development female staffed units available to assist with mental health emergency cases involving a female patient were prioritised as an action plan.
- AP 3: Secondary to this, a longer-term goal was to expand these units to encompass a dedicated mental health emergency crisis response team, to assist with cases in the community experiencing an acute episode and who are known with a significant mental health history. The crisis response team was to be developed in conjunction with the broader hospital-based mental health services to provide specialist staff and direct access to specialist facilities.
- AP 4: Development and implementation a dedicated mental health emergency clinical practice guideline (CPG), with a clear focus on patient assessment, management, and disposition. As a subset of this plan, would be the introduction of safe sedation scoring tool to provide CCPs with a guide on when and how to provide emergency sedation during a community based mental health emergency (Supplementary material 2 and 3).
- AP 5: Development and implementation a continued professional development program focusing on patient assessment and management for mental health emergencies, to be developed in conjunction with the broader specialist hospital-based mental health services.
- AP 6: Development of formal service level agreements and policies with key partner agencies
- AP 7: Develop and implementation of an activity dashboard to facilitate regular audit of the quality and performance of mental health emergencies serviced by HMCAS. This was in part created out of the data analysis for the cross-sectional study, again with a strong focus on the subset of patients requiring prehospital sedation (Figure 2). Secondary to this was the development of a “safe-sedation” clinical bundle quality indicator to be implemented for patient safety monitoring purposes in conjunction with the activity dashboard (Table 4).

**Table 4:**
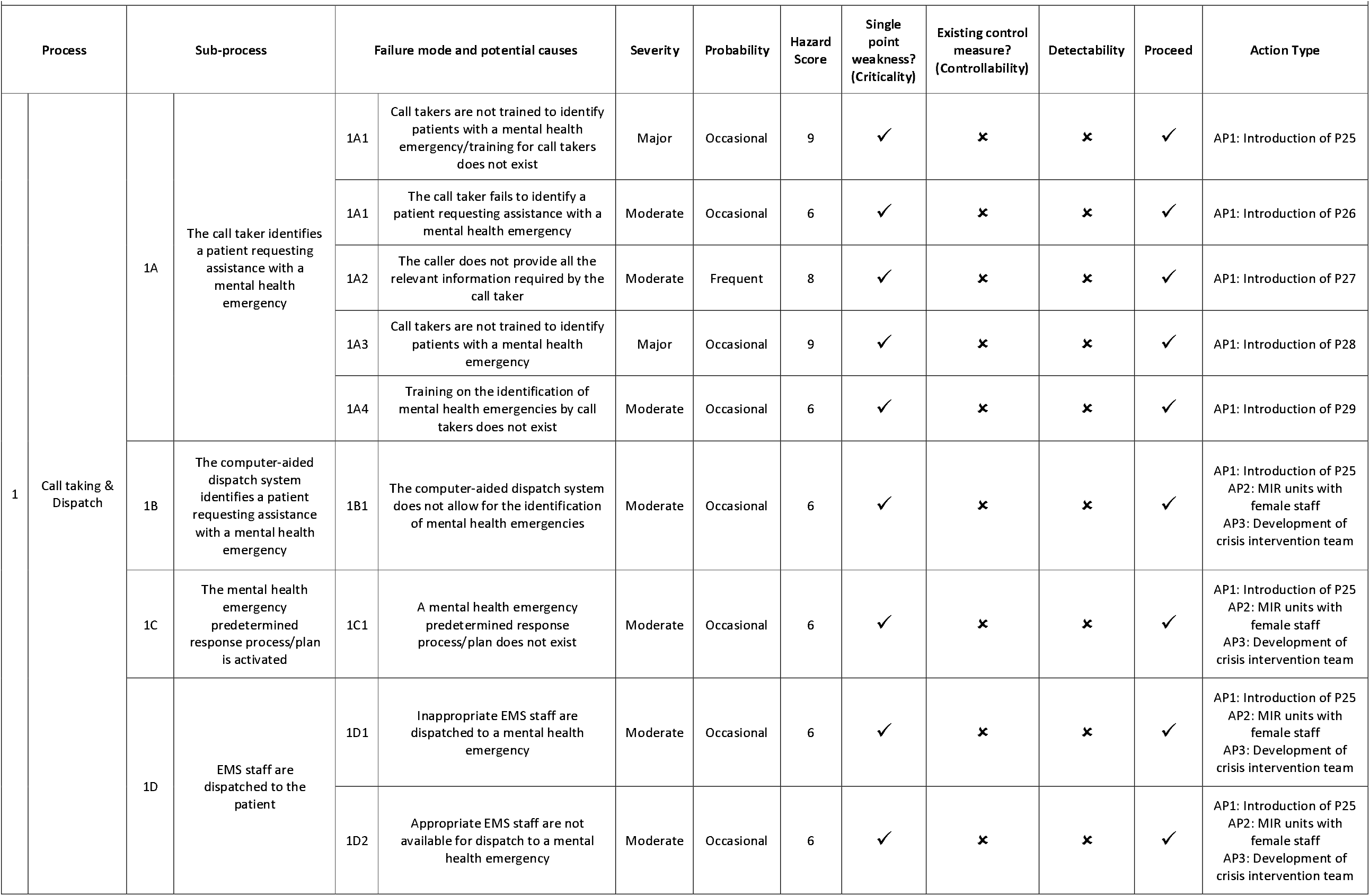
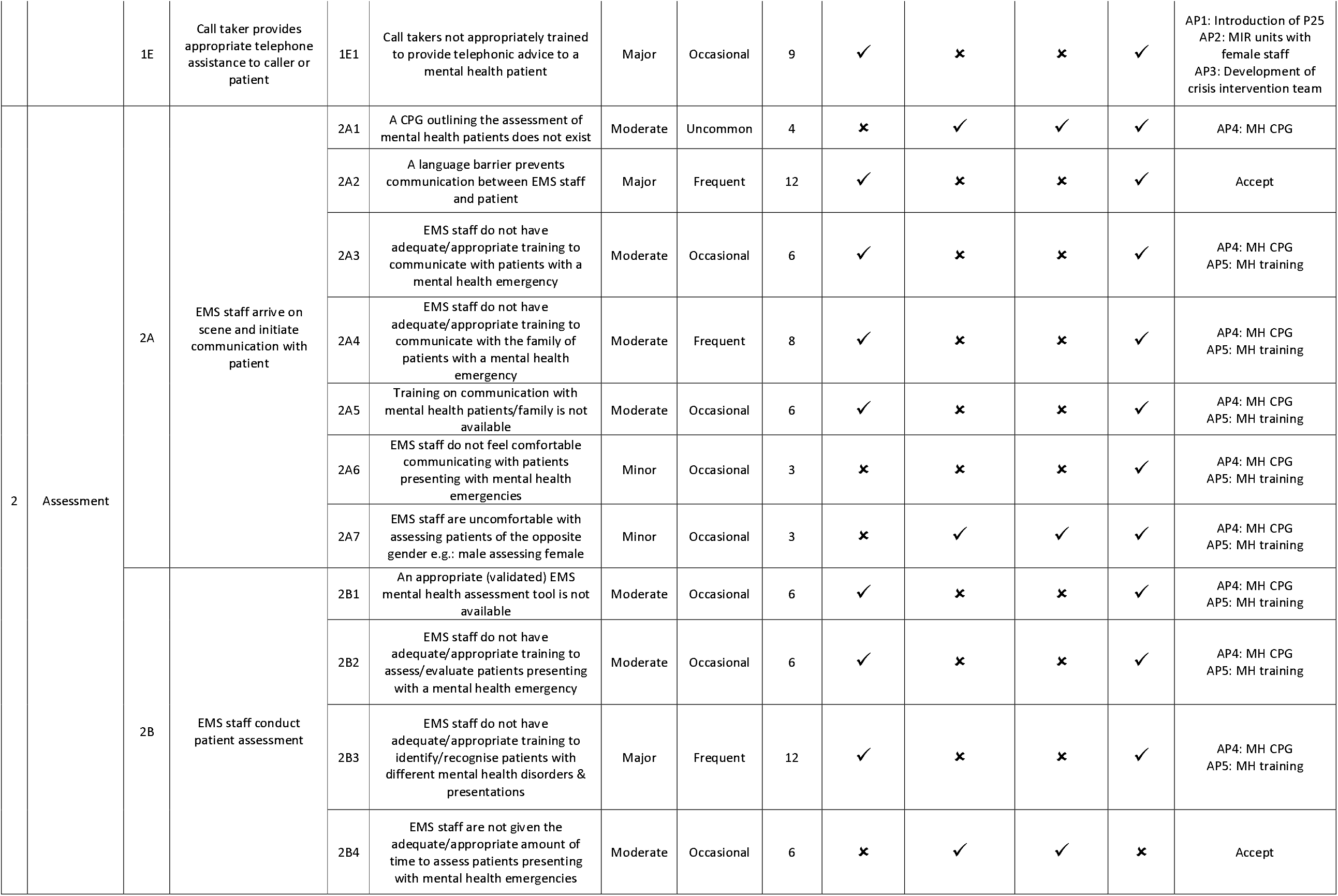
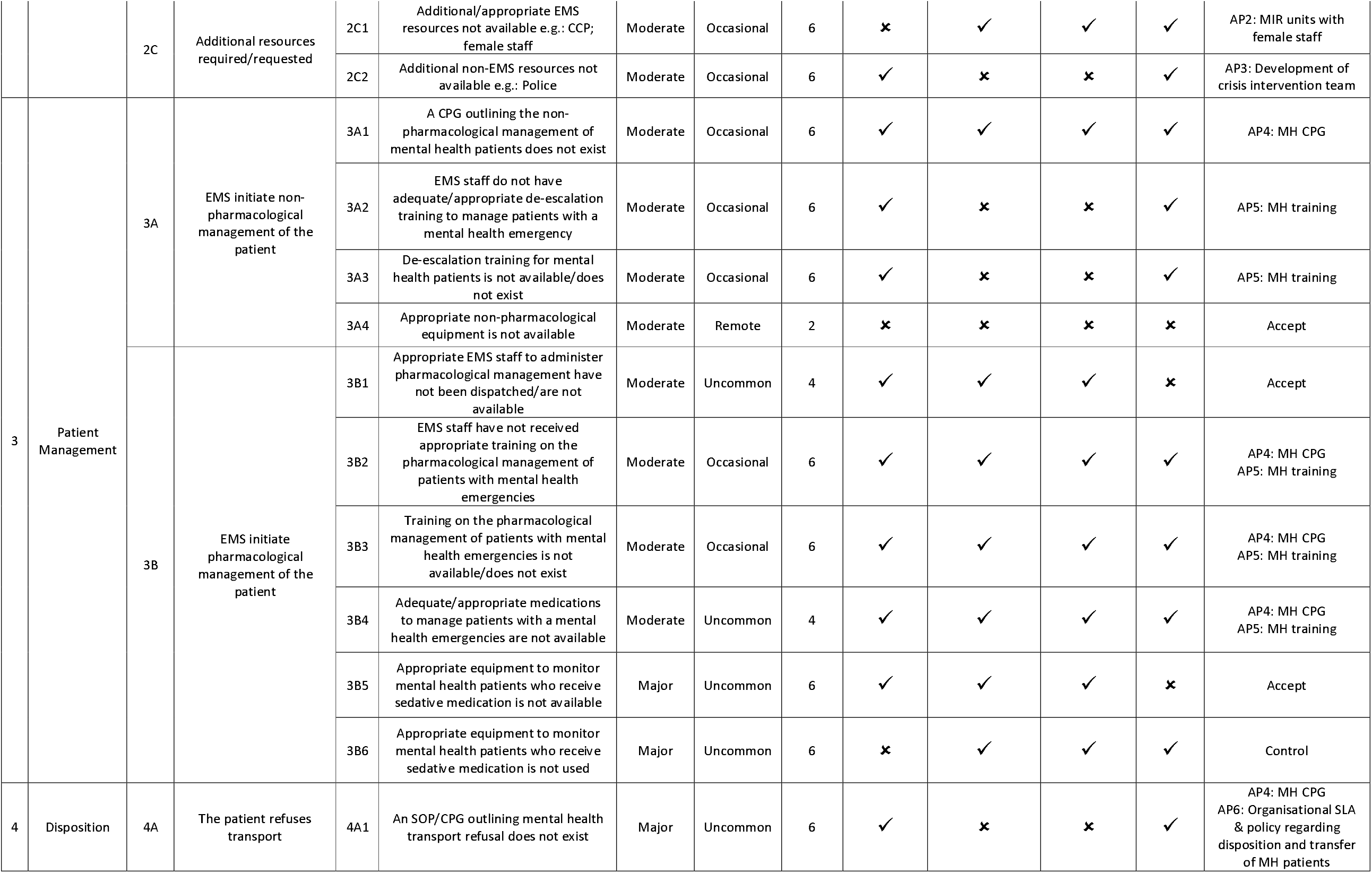
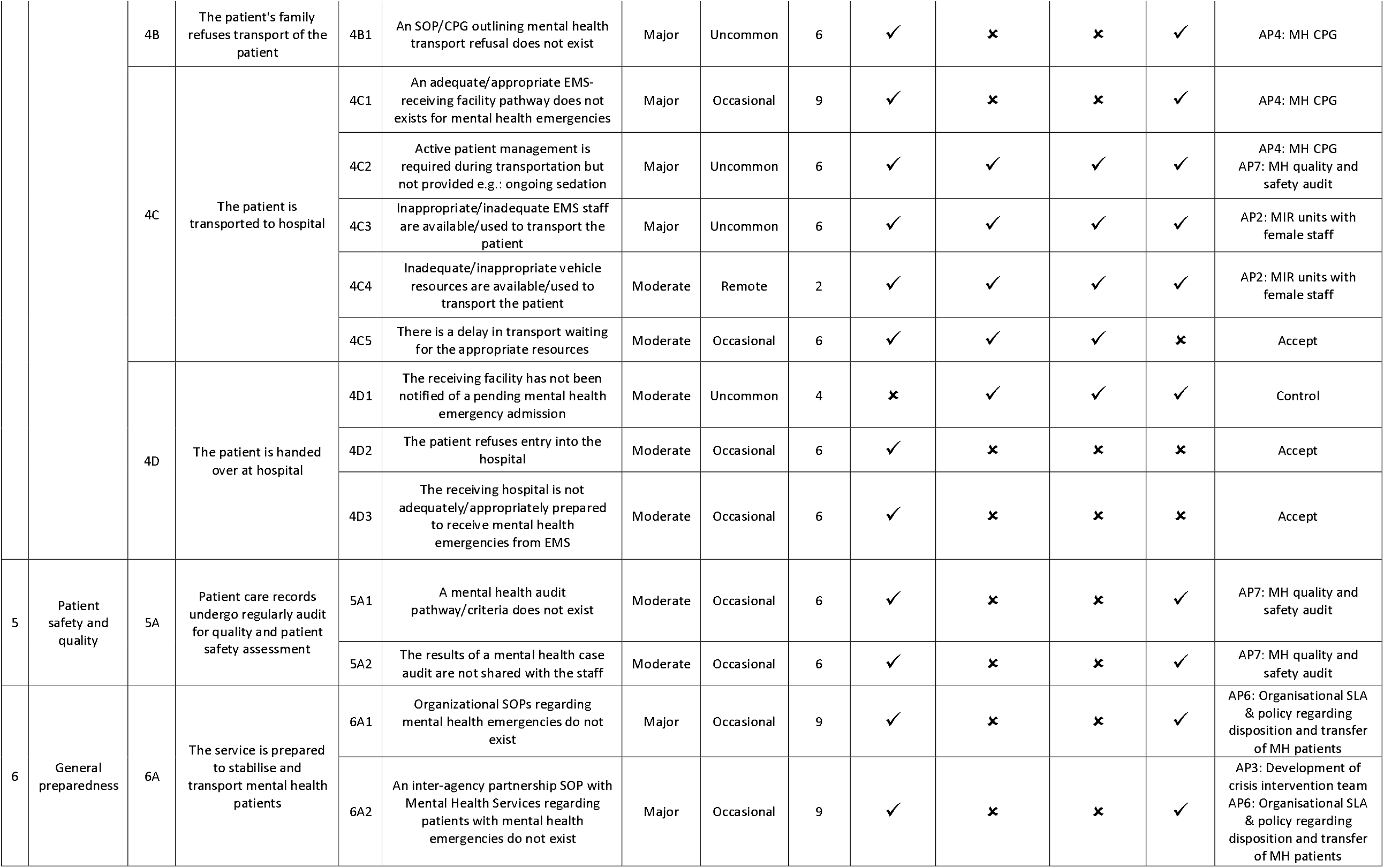
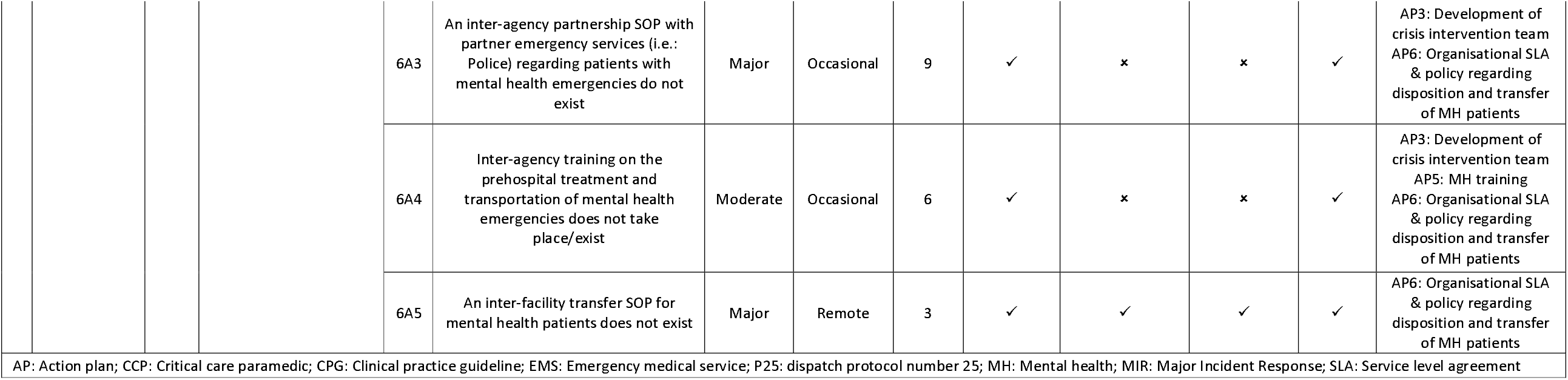
Hazard identification and action

**Table 4:** Safe sedation quality indicator data components

| Abbreviated Name | Safe sedation bundle |
| --- | --- |
| **Definition** | Patients with a provisional diagnosis of mental health emergency who received all components of the safe sedation bundle |
| **Category** | Clinical |
| **Subcategory** | Mental health |
| **Clinical Pathway/Service Pathway** | Clinical > Mental health > Safe sedation bundle |
| **Measure Type** | Process bundle |
| **Target Population** | Adult patients<br>Medical patients |
| **Unit of Analysis** | Clinician<br>Shift<br>Base<br>Service |
| **Numerator Statement** | Total number of patients with a provisional diagnosis of mental health emergency who received all components of the safe sedation bundle:<br>- Continuous 4 lead ECG monitoring<br>- Continuous SpO2 monitoring<br>- Continuous EtCO2 monitoring<br>- Continuous BP monitoring |
| **Denominator Statement** | Total number of patients with a provisional diagnosis of mental health emergency who were administered sedative medication |
| **Case Mix/Risk Adjustment** | Primary (Community) calls<br>Gender |
| **Exclusion Criteria** | Paediatric patients<br>Insufficient reporting data<br>Inter-facility transfer cases |
| **Measure Calculation** | Numerator/Denominator X 100 = % |
| **Numerical Reporting Format** | Percentage (%) |
| **Graphical Reporting Format** | Run chart (min 10 data points)<br>SPC chart (min 20 data points) |
| **Reported Indicator** | % Patients with a provisional diagnosis of mental health emergency who received all components of the safe sedation bundle |
| **Data Source** | Patient Report Form |
| **Suggested Reporting Period** | Monthly<br>Quarterly<br>Annually |
| **Recommended Review Period** | Three years |

## DISCUSSION

The utilisation of EMS for community based mental health emergencies in this study was significant, yet similar to that observed in the albeit limited literature (13). The transportation rate in the participating service, over the study period, was nearly double the activity of the more conventional case types of AMI or acute stroke. Despite this, there was unequal operational focus and clinical guidance for mental health emergencies when compared to these traditional case types. Care pathways into the broader health system in the local setting are well established, and equally common place the world over for AMI and stroke. Similarly, treatment practice guidelines are equally well established for each of these case types, yet despite evidence of greater activity, these processes are less well developed for mental health emergencies.

We, similarly, identified a high proportion of both female and younger patients transported for a mental health emergency, again echoing that seen in the literature (4-7). This presents a unique potential challenge for EMS service delivery, given the importance of communication with this patient group, and the role of the attending clinician in facilitating patient transportation through appropriate communication as a first line approach. Consequently, the role and impact of appropriate staffing is further emphasised when considering the utilisation of EMS for this patient group primarily from a communications perspective.

The most significant threat to patient safety, however, was the notable proportion of patients who received prehospital administered sedation. Further to this, utilisation of a variety of agents, potentially inappropriate agents, the use of multiple types in a single patient, and the administration of repeat doses all significantly increase the potential for an adverse drug event. In the event of a patient experiencing an acute episode, there is the additional physical threat to patient and practitioner safety in the administration of a sedative, if conducted by ill prepared or poorly trained clinicians.

In attempt to develop a care pathway focused on mental health emergencies, several significant, strategic-level hazards/failure points were identified amongst the conventional core processes within EMS, including call taking, patient assessment, management and disposition. Consequently, the focus for improvement was on conventional subprocesses within each of these that exist for the more conventional call types. Within the call taking and dispatch process, significant progress had been made on improving the identification of potential mental health emergency cases by the vendor of the computer aided dispatch program used by HMCAS. This represented the perfect opportunity to bring meaningful change towards improving the call taking process within the service and as such was incorporated as a key action item, and one that could be relatively easy incorporated into the service. The largest deficit in the service, however, was regarding the lack of clear clinical guidance towards the evaluation and management of mental health emergencies. Anecdotal feedback from frontline crew highlighted the importance of the ability to identify those patients who presented as a threat both to themselves as well as the EMS crew, as the greatest priority, followed by the ability to deescalate such situations, and as a last resort, guidance on when and how to safely provide chemical restraint for these patients in the form of sedation. Of interest to note, an underlying theme in feedback from frontline crew, that was echoed in the cross-sectional study, was the ability to address and the manage the situation when female patients presented with a mental health emergency, given the perceived high proportion of these patients that were encountered by staff. Consequently, all of these points formed the basis for the development of a focused mental health emergency CPG, in the development of a continued professional development program that would be provided to all staff into the future, and on the creation of specialised female staffed response units for use in mental health emergencies.

Lastly, amongst the panel in particular, the importance of quality, safety, and allied service cooperation were highlighted as important ancillary components of a mental health care pathway, if it were to be considered truly comprehensive. While these formed the lesser part of the process/subprocess identification and action item development, they nonetheless have an important role to play in ensuring appropriate governance towards a successful service offering for mental health patients by EMS. Towards this, the development of a new, focused mental health CPG was perceived to be an important first step towards improving the quality and safety of care of mental health emergencies. Secondary to this were the development of an activity monitoring dashboard and “safe-sedation” clinical quality bundle to monitor and guide the pathway development and improvement initiatives into the future.

## CONCLUSION

The burden of mental health emergencies on the provision of prehospital emergency medical care is significant, with the likely potential to increase into the future. Despite this, there was unequal focus on the development of care pathways and clinical guidance for this patient cohort compared with the more “traditional” case types serviced by EMS, such as MI and Stroke. Consequently, we mapped out a generic process for the service of a prehospital mental health emergency. Several potential points of failure and hazards were identified within this process. To counter these, we identified several strategic-level action plans in order to mitigate these hazards and improve the delivery of care for these patients in the community.

## Supporting information

Supplemental figure 1: Qatar population pyramid

Supplemental material 2: Acute behavioural disturbance clinical practice guideline

Supplemental material 3: Metal assessment guideline

## Data Availability

All data produced in the present work are contained in the manuscript

## DECLARATIONS

### Availability of data and material

Data can be made available upon reasonable request to the corresponding author

### Code availability

N/A

### Ethics approval

Ethical approval to conduct the study was granted by the Medical Research Centre of the Hamad Medical Corporation, Qatar (MRC-01-20-662). All study procedures and processes were conducted in accordance with that described in the ethics application and the guidelines and regulations outlined by the local medical research council.

### Author contributions

IH, NC, LAS, RO conceptualised the study aim and objectives; IH collected and analysed the data; all authors contributed equally towards the final draft of the manuscript

