## Supplemental figure 1: Qatar population pyramid for "Understanding the burden and mitigating risks in the utilisation of the Emergency Medical Services in the management of community mental health emergencies"

**Supplementary material 1: Qatar population pyramid**

Gomeseria, Ronald. (2020). "Landscape Structure and Implications for Sustainable Environmental

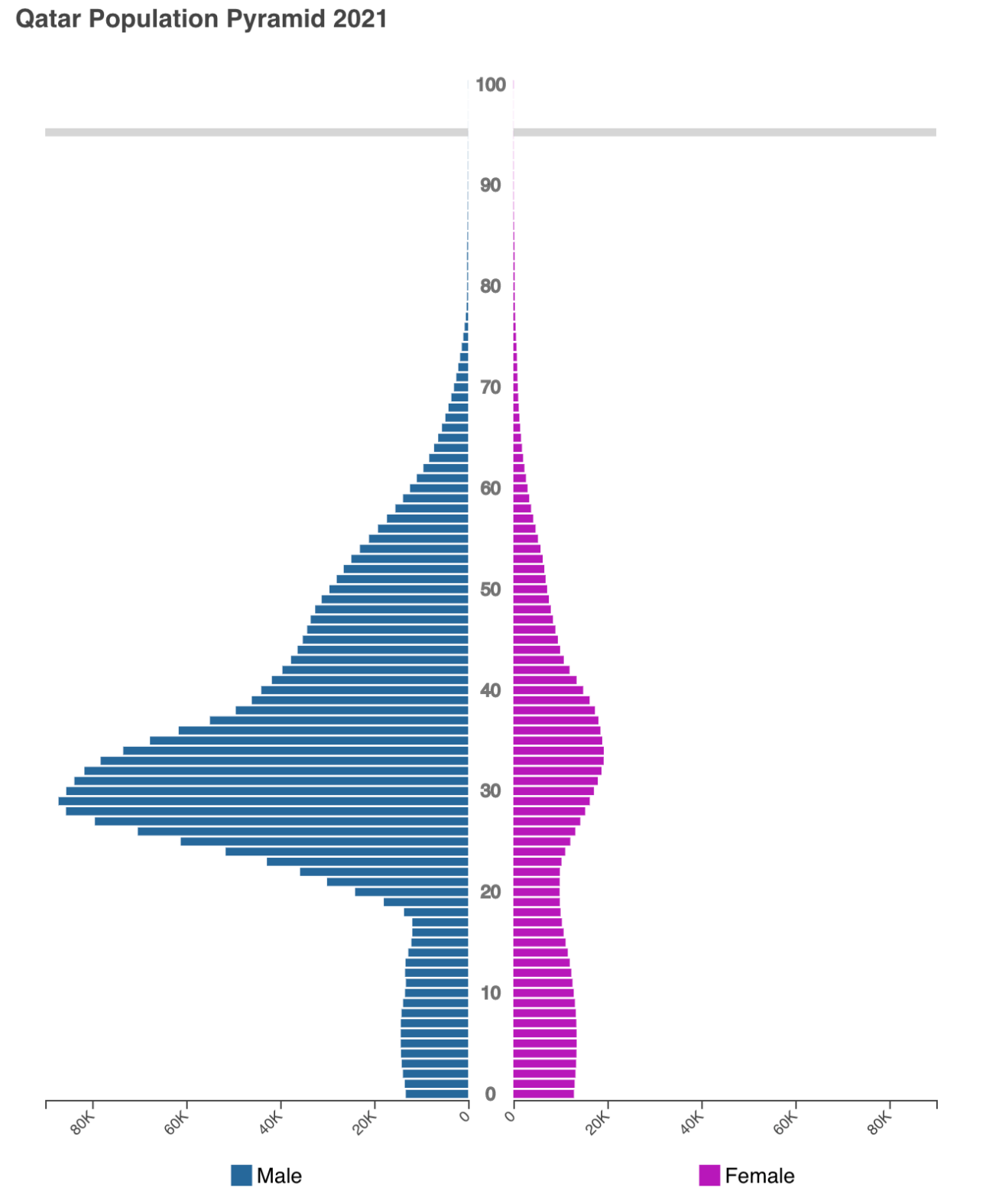
Management". 10.17605/OSF.IO/JXSK5.
