## Supplemental material 2: Acute behavioural disturbance clinical practice guideline for "Understanding the burden and mitigating risks in the utilisation of the Emergency Medical Services in the management of community mental health emergencies"

### Supplementary material 2

### MANAGEMENT OF ACUTE BEHAVIOURAL DISTURBANCE CLINICAL PRACTICE GUIDELINE

**Suspected Acute Behavioral Disturbance (ABD)**

**YES**

**Request CCP, Supervisor and Police back up if needed.**

**DO NOT** approach if it is not safe to do so

- Attempt to de-escalate the situation.
- Rule out other potential causes of ABD (i.e. hypoglycaemia).
- Use the Sedation Assessment Tool (SAT) to determine need for sedation.

**Is Sedation Required?**

**NO**

- Sedate and monitor patient as per CPG
- Record **SAT score** on the ePCR.

Transport to hospital. Pre-notify as required.

- If a patient has been physically retrained, ensure you position them appropriately and remove the restraints as soon as safely possible.
- A SAT score of +3 or +2 is a good indication of the need for sedation.

### MANAGEMENT OF ACUTE BEHAVIOURAL DISTURBANCE CLINICAL PRACTICE GUIDELINE

Patients presenting with Acute Behavioural Disturbance (ABD) pose a significant risk to themselves and the healthcare practitioners treating them. Special considerations relating to assessment, management and safety should be considered by healthcare practitioners when caring for patients with ABD.

**ASSESSMENT CONSIDERATIONS**

The causes of ABD are often multifactorial and include mental illness, intoxication with drugs and/or alcohol and organic illness such as hypoglycaemia. ABD can be classified in four general categories:

- *Psychiatric disorders –* schizophrenia, bipolar, PTSD, psychosis.
- *Substance related –* psychostimulants, cocaine, ketamine, LSD, cannabis, alcohol.
- *Organic disorders –* hypoglycaemia, sepsis, hypoxia, head injury, dementia.
- *Situational –* grief, overwhelming stress.

**Common ABD presentations include**:

- Panic
- Agitation
- Anxiety
- Delusions
- Hallucinations
- Thought disorders

When assessing and managing a patient with ABD, attempt to create a safe environment for both the healthcare practitioners and the patient. Be aware that ABD patients can be impulsive, unpredictable and a risk to themselves or others, requiring emergent treatment and management. Certain triggers may produce sudden aggression. In such cases, withdraw to safety and request supervisor, CCP and police assistance. Assessing for the cause of the ABD may sometimes only be possible after immediate management of the behavioural disturbance.

**Clues increasing likelihood of organic aetiology:**

- > 40 years of age with first presentation of psychosis or altered mental state
- Disorientation/altered LOC
- Altered vital signs
- Visual, tactile, or olfactory hallucinations
- Sudden onset
- Fluctuating conscious state

**MANAGEMENT RECOMMENDATIONS**

Management of ABD should focus on managing the possible cause of the ABD, using appropriate strategies to de-escalate the situation, and if required, safely sedating the patient. Treat reversible causes.

- Only approach the patient if it is safe to do so
- Attempt to de-escalate the situation

**Strategies for de-escalation:**

- Approach the situation with the right attitude and maintain self-control.
- Communicate non-aggression using your voice and appropriate body language.
- Match energy levels by responding appropriately and professionally to the situation. Use ‘voice for occasion’.
- Empathise and listen actively. Empathy can help defuse a conflict/ aggressive situation.
- Focus on the issue at hand. Help the patient focus on how to solve their problem.
- Consider pharmacological restraint (sedation) should other techniques fail.
- Sedation Assessment Tool (SAT) – the purpose of the SAT is to determine a patient’s level of agitation and response to medication and level of sedation.
- A SAT score of +2 or +3 is a good predictor of the need for sedation.
- Administer appropriate sedation as per CPG
- Ensure that you document the SAT score on your EPCR both prior to and post sedation.
- Transport to the nearest most appropriate healthcare facility.
- Notify CC: Emergency in order for appropriate arrangements to be made at the receiving facility.

**SEDATION ASSESSMENT TOOL (SAT)**

- Use the patients’ speech and responsiveness to determine SAT score (See table below).
- Allocate the appropriate SAT score (+3 to -3) by determining the highest-ranking value (for example, a patient displaying very anxious and agitated behavior but who speaks normally with receive a score of +2).
- Monitor patient closely following sedation using minimum monitoring requirements as per CPG.
- See SAT below.

| **SCORE** | **RESPONSIVENESS** | **SPEECH** |
| --- | --- | --- |
| +3 | Combative, violent, out of control | Continual loud outbursts |
| +2 | Very anxious and restless | Loud outbursts |
| +1 | Anxious and restless | Normal |
| 0 | Responds easily to name, speaks in normal tone | Normal |
| -1 | Responds only if name called loudly | Slurring or prominent slowing |
| -2 | Responds only to physical stimulation | Few recognizable words |
| -3 | No response | Nil |

**THE PHYSICALLY RESTRAINED PATIENT**

- As described above, the use of simple reassurance, verbal de-escalation and pharmacological restrain are to be used preferentially to avoid the use of or minimize the duration of physical restraint.
- If physical restrain is required, the least restrictive and least forceful options available should be used that do not result in pain or harm.
- If mechanical restrains are placed by police (i.e. handcuffs), the police should always be present with the patient.
- Any physical restraints used should be able to be rapidly removed.
- Physically restrained patients should be positioned on their side, preferably with the restrains in front of their body. Do not keep these patients in the prone position.
- Restraints should be removed as soon as safely possible or once appropriately sedated.
- Restrained patients should be continuously visually monitored for signs of distress and vitally monitored as per relevant CPG.
- Contact CC: Emergency so that they can notify the receiving healthcare facility in order for appropriate arrangements to be made for the patient.

**MONITORING REQUIREMENTS**

1. Vital signs monitoring as per CPG 1.
2. Continuous monitoring of SpO2 and ECG is required if sedation is administered regardless of route or dose.
3. Continuous EtCO2 monitoring is required in all cases where sedation is administered.
4. Monitoring should be applied as soon as patient is calm enough to do so safely.
