## Supplemental material 3: Metal assessment guideline for "Understanding the burden and mitigating risks in the utilisation of the Emergency Medical Services in the management of community mental health emergencies"

### Supplementary material 3

### MENTAL ASSESSMENT CLINICAL PRACTICE GUIDELINE

A mental status assessment is designed to systematically evaluate a patient’s thought processes at a particular point in time in order to guide care. Patients presenting with behavioural abnormalities should have a mental status assessment conducted if possible. It is important to note that a mental status assessment is not used to diagnose a specific condition.

| **MENTAL STATUS ASSESSMENT** | | | |
| --- | --- | --- | --- |
| - Patients presenting with a behavioural abnormality should be assessed in an attempt to identify the cause of the presenting signs and symptoms. Approach these patients in a compassionate, non-threatening manner. - Exclude and/or manage causes of abnormal behaviour where possible. - Only attempt to treat the patient if it is safe to do so. - Use the mental status assessment guide to conduct a patient assessment. The assessment should be conducted in a highly respectful and empathetic manner. Do not be disrespectful, judgemental, or interrogatory as this will most likely worsen the circumstances. - Be cognisant that different cultures hold different beliefs and view mental illness differently. Some may be accepting whereas others may see it as taboo. | | | |
| **MENTAL STATUS ASSESSMENT GUIDE** | | | |
| ***Appearance*** | - Grooming - Posture - Build - Clothing - Cleanliness | ***Thought form*** | - Amount - Rate - Derailment - Flight of ideas |
| ***Behaviour*** | - Eye contact - Mannerisms - Gait - Activity level | ***Thought content*** | - Disturbances - Delusions - Suicidal - Obsessions |
| ***Speech*** | - Rate - Volume - Pitch - Tone - Flow - Pressure | ***Perception*** | - Illusions - Thought insertion - Broadcasting - Hallucinations: - Auditory, olfactory, tactile, visual, gustatory |
| ***Mood*** | Emotion described as:   - Anxious - Depressed - Cheerful | ***Insight & judgement*** | - Cognition - Illness - Understanding - Cause & effect |
| ***Affect*** | Emotion observed as:   - Restrictive - Blunted - Labile |  |  |
